# Model for evaluating cost-effectiveness of surveillance testing for SARS-CoV2

**DOI:** 10.1101/2020.12.02.20242644

**Authors:** Jonathan Silver

## Abstract

Testing people without symptoms for SARS-CoV-2 followed by isolation of those who test positive could mitigate the covid-19 epidemic pending arrival of an effective vaccine. Key questions for such programs are who should be tested, how often, and when should such testing stop. Answers to these questions depend on test and population characteristics. A cost-effectiveness model that provides answers depending on user-adjustable parameter values is described. Key parameters are the value ascribed to preventing a death and the reproduction number (roughly, rate of spread) at the time surveillance testing is initiated. For current rates of spread, cost-effectiveness usually requires a value per life saved greater than $100,000 and depends critically on the extent and frequency of testing.

## Introduction

With renewed emphasis on testing for SARS-CoV2 (1) and the emergence of cheaper testing modalities (2), design and evaluation of testing programs are of increased importance. Since the main purpose of testing is to identify cases so that they can be isolated to prevent further spread, a critical measure is the number of secondary cases averted per dollar spent on testing. This number will be higher, all else being equal, if testing is focused on those with higher rates of infection – such as those with symptoms, contacts of those with covid-19, people who have many contacts due to their work, or who live in areas with high incidence. Once “high-yield” groups are exhausted, testing could extend to people with no identifiable high prevalence characteristics. A recent paper assessed cost-effectiveness of testing those with symptoms versus testing whole populations once or monthly, concluding that monthly testing is cost-effective under some circumstances (3). How extensive should such surveillance testing be, and how often should it be done? A model is needed to answer these questions. A little thought shows that the answers depend on many variables, such as current disease prevalence, test sensitivity, delays (if any) in isolating those who test positive, test costs, values ascribed to saving lives, etc. This paper describes a model that allows users, including those without programming skills, to easily change parameter values and compare results for a range of values. The model first calculates how much a given surveillance testing program would reduce the number of secondary cases per primary case, and then uses a “Susceptible-Infected-Removed” (SIR) epidemiological model (4) to estimate how many cases would be averted by testing over a period of time. Based on (user-modifiable) values ascribed to averting hospitalizations and deaths, the model calculates cost-effectiveness from a societal point of view, and determines the fraction of the population that should be tested and the frequency of testing to maximize cost-effectiveness. It also estimates the time at which surveillance testing ceases to be cost-effective, and the fraction of tests that are positive at this time, possibly useful as a proxy for cost-effectiveness. The model could be useful for those considering whether to implement surveillance testing, and in the design or evaluation of such programs.

### Assumptions

For simplicity, the model assumes that people with covid-19 are infectious for an average of 15 days and are equally infectious on each of these days. The duration of infectivity can be changed in the model. Consider a public health program that tests a fraction f of the population, chosen at random, every p days, with a test that has a sensitivity s (sensitivity = proportion of those with covid-19 who test positive). Further suppose that test results are available only after a potential delay so that those who test positive are isolated j days after testing, where j is an integer between 0 and 14. If j were 15 or greater, the test would not avert any cases so j>14 is not considered. Let R0 designate the average number of secondary cases per primary case in the absence of random testing. At the beginning of the pandemic, R0 was estimated to be ∼2.5, but with social distancing measures R0 has come down to ∼1.0-1.5 in the US (5).

### Number of secondary cases averted by surveillance testing

When surveillance testing is done once every 15 days or less often, the number of secondary cases prevented per primary case can be estimated as f*s*R0*(14-j)*(15-j)/(30p). A derivation is provided in the supplement. This estimate simplifies to ≈ f*s*R0/2 for j=0 and p=15 days. If only a fraction c of those tested comply with isolation, the number of secondary cases averted per primary case is reduced by a factor c. This estimate is based on multiplying the probability that testing is done on any particular day of an infected person’s infectious period, times the number of secondary cases averted if testing were done on that day, and summing over all days that result in averting cases.

If testing is done more frequently than once per infectious period, the calculation is more involved since it must consider the possibility that an infected person is not detected on the first several test dates during his/her infectious period, but then detected on a subsequent test. The number of cases averted from such late detections declines because there are fewer days left in an infectious period for isolation to have an impact. This calculation is described in more detail in the supplement.

### Optimal test fraction and frequency given budget constraint

If there were no cap on spending, the greatest reduction in spread would occur if everyone were tested (f=1) as often as possible (e.g. daily, p=1). But if an organization has a budget for random testing, f=1 and p=1 may exceed the budget; in this case what fraction f of the population should be tested, at what frequency (1/p), to provide the greatest reduction in spread? The optimum turns out to be testing once per infectious period or less often, with the fraction tested determined by how many tests the budget allows when testing is performed every p days. Mathematical details are provided in the Supplement. In other words, all combinations of f and p constrained by the budget, but with p greater than or equal to the infectious period, are equally efficient in reducing secondary cases. Testing more frequently than once per infectious period is less efficient because then some of the cases detected come from those tested more than once, resulting in more dollars being spent per case detected.

### Guideline for when confirmatory polymerase chain reaction (pcr) tests should be done for those who test positive on screening tests

Antigen tests for SARS-CoV2 recently approved by the FDA promise to be less expensive than pcr but may suffer from high false positive rates (2). False positives from non-pcr screening tests could be dealt with by doing a confirmatory pcr test on all those whose screening test is positive. From a cost-effectiveness point of view, confirmatory pcr tests should be done when the cost of doing confirmatory tests is less than the cost of isolating those who falsely test positive. An argument in the supplement shows that this is equivalent to doing confirmatory tests when the false positive rate exceeds the ratio of the cost of a pcr test to the cost of isolation. The latter might be estimated as the average value of 2 weeks of lost income or about $2000 in the US. If pcr tests cost $100, then it would be cost effective to use them to confirm positive screening tests when the false positive rate exceeds 5 percent.

### User-friendly cost-benefit model that facilitates comparing effects a range of parameter values

A more complete cost-benefit analysis takes into account testing costs, societal costs from isolations, money saved from hospitalizations averted (6), and monetized value of lives saved (7). A detailed description of the model is provided in the supplement. While potentially controversial, a ‘value per life’ is used by government agencies in other contexts, such as deciding how much to spend on highway safety infrastructure (7). This value is sometimes referred to a as the value of a statistical life; here it is designated ‘spending cap per life saved’. Values of spending cap per life saved ranging from $100,000 to $1M are considered in the following.

This model yields values for optimal fraction tested and frequency of testing that sometimes differ from those that maximally reduce R0; for example, sometimes optimal testing frequency is more than once per infectious period. This is not surprising because “extra” dollars spent on reducing R0 early can reduce the extent of the epidemic over time.

Figure 1 shows a plot from the cost-effectiveness model for cumulative net benefit in $ per capita over time, where time is measured in units of 15 days (1 infectious period). Benefit curves for different spending caps per life saved are shown in different colors. The diagonal and downward directed arrows at the top of the display permit the user to toggle among a range of values for the number of days isolation is delayed, rate of compliance with isolation, test frequency, test sensitivity, cost of screening test, R0 before surveillance testing is initiated, and fraction tested; the cumulative net benefit curve updates immediately when these values are changed. In general, the cumulative benefit rises over time, reaches a peak, and then falls as the epidemic subsides. During the rising phase, the incremental benefits from random testing exceed incremental costs. At the peak of cumulative net benefit, incremental costs just equal incremental benefits. At later times, disease prevalence is sufficiently low that surveillance testing is no longer cost effective. Thus, the time when cumulative net benefit peaks is an estimate of when surveillance testing ceases to be cost effective. At early times cumulative net benefit may be negative or show a small depression because benefits from reducing spread take time to accrue whereas testing costs are constant in time. For the parameter values indicated in the top portion of the figure, surveillance testing is marginally cost effective at a spending cap per life saved of $200,000 (blue curve). Higher values of R0 make surveillance testing cost-effective at lower values of spending cap per life saved. The value of spending cap per life saved that makes the peak of cumulative net benefit just greater than zero is effectively the net cost per life saved from surveillance testing, a measure that could be compared to costs per life saved from other interventions. The net cost per life saved from surveillance testing should be evaluated at values of fraction tested and frequency of testing that maximize cumulative net benefit.

**Figure 1.**
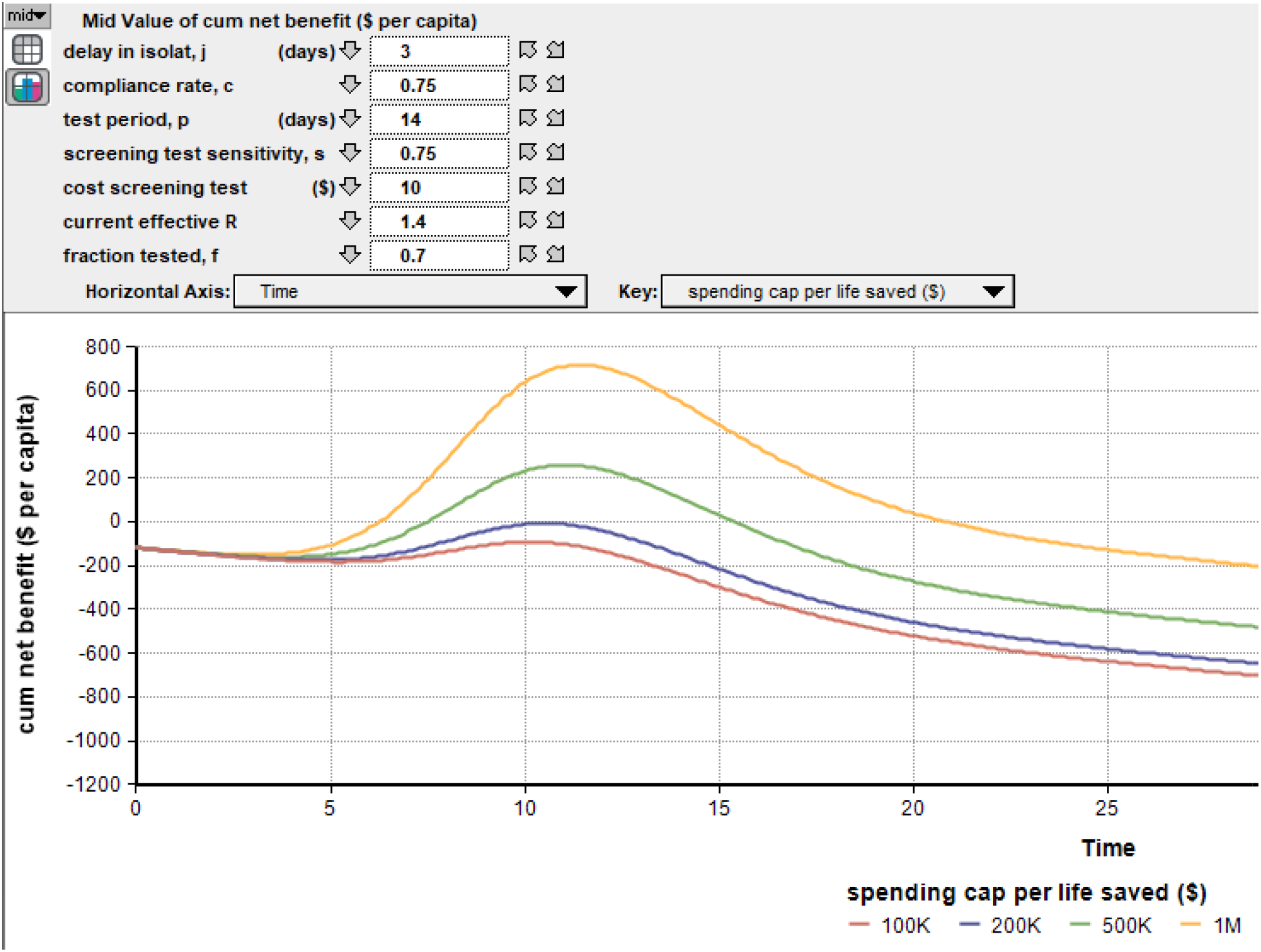
Cumulative net benefit (in $ per capita) of surveillance testing over time, for four values of spending cap per life saved ($1M, $500,000, $200,000, $100,000). Time is in units of infectious periods (∼2 weeks). Given parameter values shown at the top, surveillance testing is just cost-effective at a spending cap per life saved of $200,000. Running the program, the user can choose among a range of parameter values using the diagonal or downward-directed arrows at the top. Current effective R is R0 in the absence of surveillance testing. Other parameters that can be set by the user are shown in Supplement Figure e1.

Figure 2 is a table of the values of fraction tested f and period of testing p that maximize cumulative net benefit, and the time when net benefit is maximal, for screening tests costing $10, $30 or $100, with other parameters shown at the top. When surveillance testing is marginally cost-effective, choosing optimal values for extent of testing and frequency of testing is crucial for overall cost-effectiveness.

**Figure 2.**
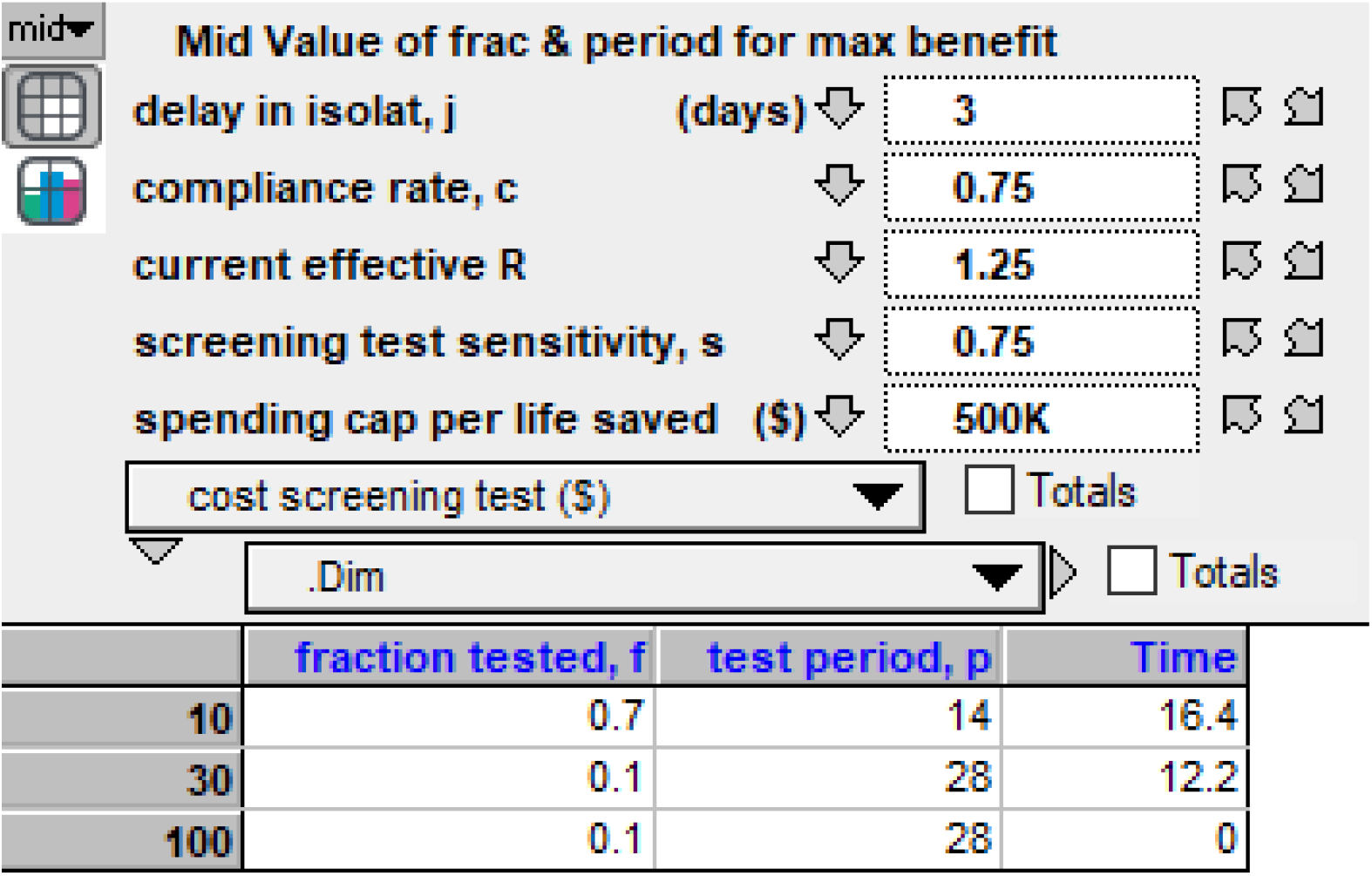
Table of values for fraction of population tested and test period that maximize cumulative net benefit, for screening test costs (rows) of $10, $30, and $100. The time when cumulative net benefit is maximal is given in the fourth column (in units of infectious periods, ∼2 weeks). Time =0 is returned when testing is not cost-effective, as indicated here when screening tests cost $100. Parameters whose values can be changed by the user using the diagonal or downward-directed arrows are shown in the upper portion of the figure. Other parameters that can be set by the user are shown in Supplement Figure e1.

Figure 3 is a plot of new infections (per tenth of an infectious period) per capita over time with, versus without, surveillance testing. In this illustration, infections decrease monotonically over time given surveillance testing (blue curve) because the starting value of R0 (1.1) was chosen to be only slightly above 1 and testing immediately reduced it to below 1.

**Figure 3.**
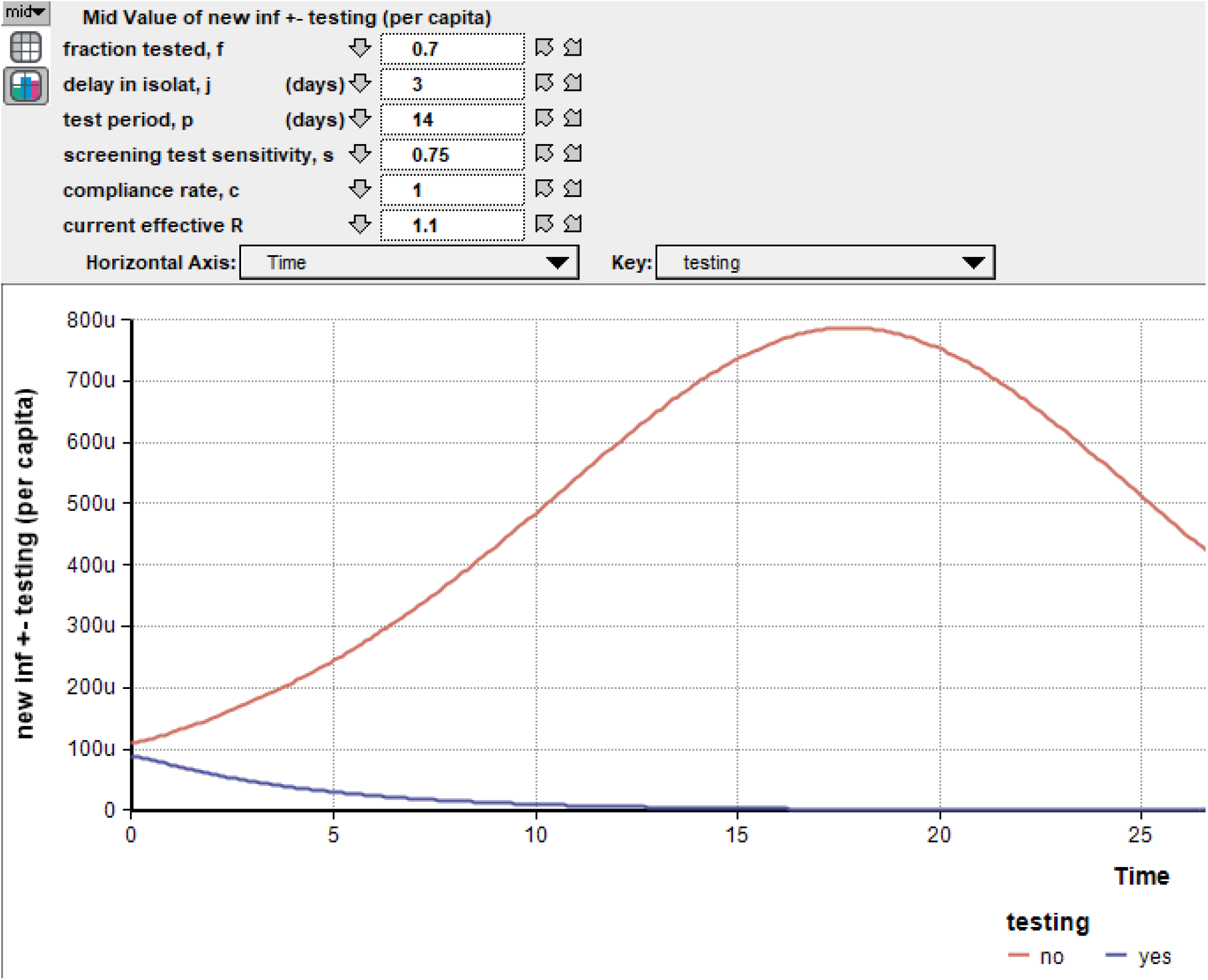
Disease incidence over time (in units of number of new cases per tenth of an infectious period per capita) with, versus without, surveillance testing. “u” on the y axis denotes cases per million people. Parameters whose values can be changed by the user using the diagonal or downward-directed arrows are shown in the upper portion of the figure. Other parameters that can be set by the user are shown in Supplement Figure e1.

Table 1 shows the fraction of tests that are positive (“critical fractions”) at the time when the model suggests surveillance testing just ceases to be cost-effective, for illustrative values of isolation delay, compliance rate, and screening test cost. In all cases, frequency of testing and fraction of population tested were chosen to maximize cost-effectiveness. Of note, sometimes the critical fractions are considerably lower than thresholds suggested for test positivity in clinical testing (8).

**Table 1.**
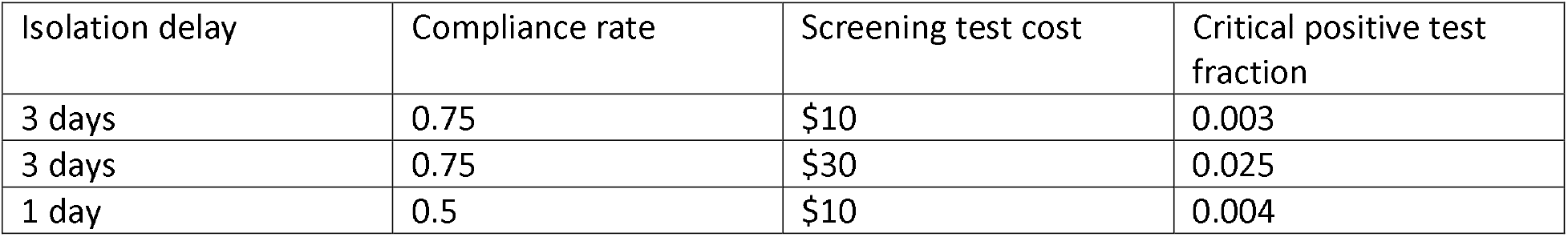
Critical positive test fraction. Fraction of screening tests that are positive when surveillance testing just ceases to be cost-effective is shown in fourth column for illustrative values of isolation delay, compliance rate and screening test cost. In all cases, spending cap per life saved was $500K, R0 was 1.25, test sensitivity was 0.75, and test frequency and fraction of population tested were chosen to maximize cost-effectiveness.

The model was implemented in Analytica 101 software, which can be downloaded for free (9). The model is free on request from the author and runs using the Analytica 101 software, allowing users to compare results from a range of parameter values. Figure e1 in the supplement shows the graphical interface where users can change parameter values, or ranges of parameter values, and select outputs for display.

### Model limitations

The model does not allow for changing the fraction tested or frequency of testing *during* a testing program; in theory, this could increase cost-effectiveness. Nor does the model account for possible changes in R0 *during* a testing program due to behavioral changes or public health measures other than surveillance testing. As a practical matter, this flexibility could be achieved by considering model results up to a time t, and then running the model for an additional period starting with revised estimates of R0 and initial parameters appropriate to time t. This could be important if, for example, vaccination during a surveillance testing program led to rapid reduction in the fraction of the population that is susceptible. The model is conservative in that it does not consider indirect economic benefits from curtailing the epidemic, such as reductions in jobs lost.

## Conclusions

A model is presented that allows rapid assessment of the impact of surveillance testing on epidemic dynamics and optimization of program variables such as extent and frequency of testing. An important feature of the model is that it gives users flexibility in choosing values for parameters that involve value judgement or political consideration (such as value per life saved), or may change over time during the epidemic (such as number of secondary cases per primary case).

## Supporting information

CHEERS checklist

## Data Availability

No data is reported in the manuscript. The Analytica model program described in the manuscript is freely available from the author.

## Acknowledgments

The author thanks Lonnie Chrisman of Lumina.com for assistance with the Analytica 101 program.

The author has no conflicts of interest, including no relationship with Lumina, the company that created the Analytica 101 software. No money was received to support this study.

