## Supplement for "Model for evaluating cost-effectiveness of surveillance testing for SARS-CoV2"

**Reduction in number of secondary cases per primary case.** The chance an infected person is detected equals f*s (the chance he/she is selected for testing times the probability the test is positive). If this person were tested on the *first* day of his/her infectivity and isolated j days later, R0*(15-j)/15 cases would be averted; if this person were tested on the *nth* day of infectivity, R0*(15-j-n)/15 cases would be averted; n can be an integer from 1 to 15-j since the number of cases averted can’t be <0. The *expected* number of cases averted equals the probability (1/p) that the test is done on the nth day of infectivity, times the number of cases averted if the test is done on the nth day, summed over permissible values of n. This sum contains the sum of integers from 1 to 14-j, which equals (14-j)(15-j)/2.

If testing is done more frequently than once per infectious period, the probability that a covid-19 case is detected in the i+1^th^ round of testing after having been missed in the previous i rounds (all rounds occurring during one infectious period) is f*s*(1-f*s)^i. If the first round of testing happened to occur on the first day of the testee’s infectious period, the number of days of potential spread averted would be 15-j-i*p, for values of j, i and p for which 15-j-i*p > 0; if it occurred on the n^th^ day of the infectious period, the number of days of potential spread averted would be 15-j-i*p-n. Multiplying by (1/p)=the probability that the first round of testing occurs on any given day of an infectious period and summing over all such days consistent with the constraint that 15-j-i*p-n >0, gives the expected number of days of potential spread averted.

**Optimal fraction tested and test frequency to maximize the number of secondary cases averted given a fixed budget for testing.** The number of tests done in a time period T>> p equals f*N*T/p where N is the population size; hence the number of tests done per capita per unit time period equals f/p. If the budget cap constrains, the number of tests that can be done per capita per unit time period = b/cpt where b is the budget per capita per unit time period and cpt is the cost per test. Equating these implies p = f*cpt/b. Substituting this value for p in the expression for expected number of cases averted makes it a function of f alone, when j, s, b and cpt are considered parameters. Maximizing the expected number of cases averted with respect to f then determines f_opt and p_opt. The result is that all values of p greater than the infectious period, with f_opt = p*b/cpt, are equivalent in maximizing reduction in cases.

**Cost-effectiveness of confirmatory pcr tests.** It would be cost-effective to do confirmatory pcr tests if the total cost of pcr tests on all those whose screening test is positive is less than the total cost of isolating false positives. If n is the number of positive screening tests, the cost from isolating false positives is n*false positive rate*cost of isolation, whereas the cost of confirming all positive screening tests is n*cost of pcr test.

**Description of more detailed model.** Cases over time are predicted from a SIR model given initial values for fraction of the population susceptible, infected, and removed (= recovered, dead or immune), and R0 before testing. Reduction in R0 due to testing, if testing is done, is calculated as described above, given values of test frequency, sensitivity, days delay in isolating those who test positive, and rate of compliance with isolation. The number of deaths and hospitalizations are calculated from the number of cases times case fatality rate (1) and hospitalization rate parameter, respectively. Monetary benefits from testing are calculated as the difference in number of deaths and hospitalizations with, versus without, testing, multiplied by values for spending cap per life saved and average hospitalization cost, respectively. Costs are the sum of costs from testing and isolation. Testing costs are number of tests done (determined by fraction of population tested and frequency of testing) times a cost per screening test, plus the number of confirmatory tests done (the sum of true positive and false positive tests) times a cost per confirmatory test. All positive screening tests were assumed to be followed by a confirmatory test. Isolation costs are the number of isolations (determined by number of true positive tests and the compliance rate) times a cost per isolation. Cumulative net benefit is the difference between monetary benefit and cost, cumulated over time. A discount rate was not used because of the short time duration considered.

Supplement. Figure e1. User interface for entering parameter values (or lists of values) for variables and selecting outputs to be displayed (cyan). Model parameters are grouped into those with single estimates (left column; these values can be changed by clicking on the orange bars and typing in new values), and parameters for which the user might want to consider a range of values (middle column; the user can change values in the list by clicking on the List, and toggle among values in the list in the output displays as shown at the tops of figures 1-3 in the main text). Parameters under a decision maker's control are shown in green bars, middle column. Double-clicking on “The model” yields a diagram of model structure with model details.

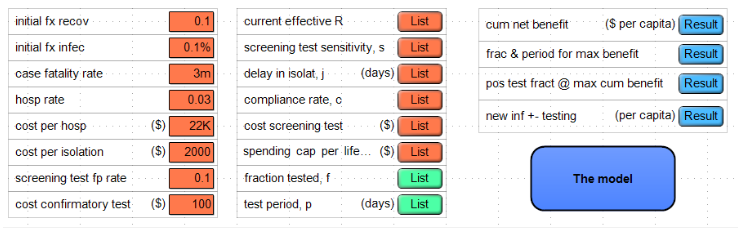

Supplement References

1. Ioannides JP. Infection fatality rate of COVID-19 inferred from seroprevalence data. Bulletin of the World Health Organization; Type: Research Article ID: BLT.20.265892 <https://www.who.int/bulletin/online_first/BLT.20.265892.pdf>
