## Supplementary material for "Model for evaluating cost-effectiveness of surveillance testing for SARS-CoV2": CHEERS checklist

### CHEERS Checklist
### Items to include when reporting economic evaluations of health interventions

The **ISPOR CHEERS Task Force Report**, *Consolidated Health Economic Evaluation Reporting Standards (CHEERS)—Explanation and Elaboration: A Report of the ISPOR Health Economic Evaluations Publication Guidelines Good Reporting Practices Task Force,* provides examples and further discussion of the 24-item CHEERS Checklist and the CHEERS Statement. It may be accessed via the *Value in Health* or via the ISPOR Health Economic Evaluation Publication Guidelines – CHEERS: Good Reporting Practices webpage: http://www.ispor.org/TaskForces/EconomicPubGuidelines.asp

| Section/item | Item No | Recommendation | Reported on page No/ line No |
|---|---|---|---|
| **Title and abstract** | | | |
| Title | 1 | Identify the study as an economic evaluation or use more specific terms such as "cost-effectiveness analysis", and describe the interventions compared. | 1/1 |
| Abstract | 2 | Provide a structured summary of objectives, perspective, setting, methods (including study design and inputs), results (including base case and uncertainty analyses), and conclusions. | 3 |
| **Introduction** | | | |
| Background and objectives | 3 | Provide an explicit statement of the broader context for the study.<br>Present the study question and its relevance for health policy or practice decisions. | 4 |
| **Methods** | | | |
| Target population and subgroups | 4 | Describe characteristics of the base case population and subgroups analysed, including why they were chosen. | NA |
| Setting and location | 5 | State relevant aspects of the system(s) in which the decision(s) need(s) to be made. | 4/11 |
| Study perspective | 6 | Describe the perspective of the study and relate this to the costs being evaluated. | 4/21 ; 19/10-23 |
| Comparators | 7 | Describe the interventions or strategies being compared and state why they were chosen. | |
| Time horizon | 8 | State the time horizon(s) over which costs and consequences are being evaluated and say why appropriate. | 8/5-18 |
| Discount rate | 9 | Report the choice of discount rate(s) used for costs and outcomes and say why appropriate. | 20/1 |
| Choice of health outcomes | 10 | Describe what outcomes were used as the measure(s) of benefit in the evaluation and their relevance for the type of analysis performed. | 19/10-23 |
| Measurement of effectiveness | 11a | *Single study-based estimates:* Describe fully the design features of the single effectiveness study and why the single study was a sufficient source of clinical effectiveness data. | NA |

| | | | |
|---|---|---|---|
| | 11b | *Synthesis-based estimates:* Describe fully the methods used for identification of included studies and synthesis of clinical effectiveness data. | NA |
| Measurement and valuation of preference based outcomes | 12 | If applicable, describe the population and methods used to elicit preferences for outcomes. | NA |
| Estimating resources and costs | 13a | *Single study-based economic evaluation:* Describe approaches used to estimate resource use associated with the alternative interventions. Describe primary or secondary research methods for valuing each resource item in terms of its unit cost. Describe any adjustments made to approximate to opportunity costs. | NA, [illegible] |
| | 13b | *Model-based economic evaluation:* Describe approaches and data sources used to estimate resource use associated with model health states. Describe primary or secondary research methods for valuing each resource item in terms of its unit cost. Describe any adjustments made to approximate to opportunity costs. | Refs 5,6,7<br>Supplement ref 1 |
| Currency, price date, and conversion | 14 | Report the dates of the estimated resource quantities and unit costs. Describe methods for adjusting estimated unit costs to the year of reported costs if necessary. Describe methods for converting costs into a common currency base and the exchange rate. | NA |
| Choice of model | 15 | Describe and give reasons for the specific type of decision-analytical model used. Providing a figure to show model structure is strongly recommended. | model structure figure shown in program available from author<br>See supplement fig e1 |
| Assumptions | 16 | Describe all structural or other assumptions underpinning the decision-analytical model. | 19/10-23 |
| Analytical methods | 17 | Describe all analytical methods supporting the evaluation. This could include methods for dealing with skewed, missing, or censored data; extrapolation methods; methods for pooling data; approaches to validate or make adjustments (such as half cycle corrections) to a model; and methods for handling population heterogeneity and uncertainty. | 19/10·23 |
| **Results** | | | |
| Study parameters | 18 | Report the values, ranges, references, and, if used, probability distributions for all parameters. Report reasons or sources for distributions used to represent uncertainty where appropriate. Providing a table to show the input values is strongly recommended. | NA |
| Incremental costs and outcomes | 19 | For each intervention, report mean values for the main categories of estimated costs and outcomes of interest, as well as mean differences between the comparator groups. If applicable, report incremental cost-effectiveness ratios. | Figures 1–4 |
| Characterising uncertainty | 20a | *Single study-based economic evaluation:* Describe the effects of sampling uncertainty for the estimated incremental cost and incremental effectiveness parameters, together with the impact | NA |

| | | | |
|---|---|---|---|
| | | of methodological assumptions (such as discount rate, study perspective). | |
| | 20b | *Model-based economic evaluation:* Describe the effects on the results of uncertainty for all input parameters, and uncertainty related to the structure of the model and assumptions. | Figs 1-4 |
| Characterising heterogeneity | 21 | If applicable, report differences in costs, outcomes, or cost-effectiveness that can be explained by variations between subgroups of patients with different baseline characteristics or other observed variability in effects that are not reducible by more information. | NA |
| **Discussion** | | | |
| Study findings, limitations, generalisability, and current knowledge | 22 | Summarise key study findings and describe how they support the conclusions reached. Discuss limitations and the generalisability of the findings and how the findings fit with current knowledge. | 10 |
| **Other** | | | |
| Source of funding | 23 | Describe how the study was funded and the role of the funder in the identification, design, conduct, and reporting of the analysis. Describe other non-monetary sources of support. | 13 |
| Conflicts of interest | 24 | Describe any potential for conflict of interest of study contributors in accordance with journal policy. In the absence of a journal policy, we recommend authors comply with International Committee of Medical Journal Editors recommendations. | 13 |

For consistency, the CHEERS Statement checklist format is based on the format of the CONSORT statement checklist

The **ISPOR CHEERS Task Force Report** provides examples and further discussion of the 24-item CHEERS Checklist and the CHEERS Statement. It may be accessed via the *Value in Health* link or via the ISPOR Health Economic Evaluation Publication Guidelines – CHEERS: Good Reporting Practices webpage: http://www.ispor.org/TaskForces/EconomicPubGuidelines.asp

The citation for the CHEERS Task Force Report is:
Husereau D, Drummond M, Petrou S, et al. Consolidated health economic evaluation reporting standards (CHEERS)—Explanation and elaboration: A report of the ISPOR health economic evaluations publication guidelines good reporting practices task force. Value Health 2013;16:231-50.
